# High Prevalence of Molecular Markers Associated with Artemisinin, Sulphadoxine and Pyrimethamine Resistance in Northern Namibia

**DOI:** 10.1101/2025.01.09.25320247

**Authors:** Lydia Eloff, Andrés Aranda-Díaz, Isobel Routledge, Amy Wesolowski, Mukosha Chisenga, Brighton Mangena, John Chimumbwa, Chadwick Sikaala, Petrina Uusiku, Stark Katokele, Jaishree Raman, Jennifer Smith, Davis R. Mumbengegwi

## Abstract

Artemisinin-based combination therapies are a cornerstone of Namibia’s efforts to eliminate malaria, which has had an over 90% reduction in cases since their introduction in 2005. However, their efficacy has not been routinely monitored, with malaria outbreaks regularly reported since 2016. The recent emergence of artemisinin partial resistance in Africa has highlighted the role of malaria molecular surveillance in complementing efficacy studies. This cross-sectional genomic surveillance study was nested within Namibia’s routine surveillance system and aimed to determine the prevalence of antimalarial drug resistance markers in northern Namibia. Dried blood spots (DBS) and epidemiological data were collected from confirmed *Plasmodium falciparum* cases presenting at health facilities in the highest malaria-burden regions (Zambezi, Kavango East, Kavango West, Ohangwena, and Omusati) from April to September 2023. 12 genes associated with resistance to 7 antimalarial drugs were genotyped from 264 DBS using multiplexed targeted amplicon sequencing. Multiple *pfk13* mutations associated with artemisinin partial resistance were identified: the P441L candidate marker was the most abundant, at 33.2%, and the validated markers P574L and A675V were observed at 1.2%. The chloroquine resistance marker *pfcrt* CVIET haplotype was observed at 1%, while the *pfmdr1* N86 genotype, selected by lumefantrine use, was found in all samples. Although sulphadoxine-pyrimethamine is not used in Namibia, a high proportion of sulphadoxine-pyrimethamine resistance-associated mutations in the *pfdhps* and *pfdhfr* genes were observed. This study underscores the need for routine genomic surveillance to monitor emerging drug resistance markers and calls for further research to define their clinical implications.

## Introduction

Namibia, a low malaria transmission southern African country targeting elimination, still has 1.7 million people (68% of the population) at risk of contracting the disease.^1^ Despite substantial progress in reducing cases from 538,512 in 2001 to 3,404 in 2019, progress has plateaued, with outbreaks regularly occurring, as evidenced by the 12,993 cases reported in 2023 (Ministry of Health and Social Services, personal communication).^2,3^ Over the last decade, malaria transmission has shifted from north-central Namibia to the northeastern regions of Kavango East, Kavango West, and Zambezi.^4^ Challenges to malaria elimination include: behavioral changes in *Anopheline* vectors; a concentration of cases in high-risk groups such as young men and travelers; cross-border population movement driving importation; and asymptomatic and low parasitemia infections.^5–8^ While *Plasmodium falciparum* accounts for the majority of the infections, other *Plasmodium* species pose unique challenges due to their distinct biology and transmission dynamics.^9^ Finally, the potential emergence and spread of antimalarial drug resistance threaten the efficacy of current treatment regimens, underscoring the need for sustained vigilance.^10^

Chloroquine resistance was first detected in Namibia in 1984, with increasing resistance leading to a policy shift in 2005.^11^ The artemisinin-based combination therapies (ACT), artemether-lumefantrine and dihydroartemisinin-piperaquine, replaced chloroquine as first- and second-line malaria treatment, respectively.^12^ Growing reports of artemisinin partial resistance in Sub-Saharan Africa underscore the urgent need for ongoing surveillance and efficacy monitoring.^13^ Without routine surveillance for antimalarial resistance, early warning signs may be missed, risking the increased selection for and spread of resistant parasites during outbreaks, undermining Namibia’s elimination efforts.

Antimalarial efficacy is typically monitored through therapeutic efficacy studies (TES), which are resource-intensive and impractical in low-transmission settings with highly mobile populations due to difficulties in achieving target sample sizes.^14^ Therefore, in settings with very low transmission, the World Health Organization has recommended adaptations to the TES protocol or the implementation of integrated drug efficacy surveillance (iDES), along with enhanced molecular surveillance.^15^ Routine molecular surveillance of resistance markers, including cost-effective genomic methods, can provide early detection of emerging drug resistance.^15–17^ Single nucleotide polymorphisms (SNPs) in the propeller domain of PfK13 (coded by the *pfk13* gene) have been found to be associated with delayed parasite clearance in vitro and in vivo after artemisinin treatment, a sign of artemisinin partial resistance.^18,19^ These SNPs include ‘candidate’ (significant clinical or laboratory evidence for the association) and ‘validated’ (both clinical and laboratory evidence) markers.^14^ Multiple markers of artemisinin partial resistance have recently independently emerged in Sub-Saharan Africa, including P441L, C469Y/F, R561H, and A675V in East Africa (Uganda, Rwanda, Tanzania), and R622I in the Horn of Africa (Ethiopia, Eritrea).^20–25^

Molecular markers associated with resistance to other antimalarial drugs have been identified and include SNPs associated with resistance to sulphadoxine (in the dihydropteroate synthesase, *pfdhps*, gene), pyrimethamine (in the dihydrofolate reductase, *pfdhfr*, gene), and chloroquine (in the chloroquine resistance transporter, *pfcrt*, gene).^26^ These markers are important for ACT partner and non-front-line drug resistance assessment. Mutations in the multidrug resistance 1 (*pfmdr1*) gene modulate chloroquine resistance and are differentially selected for by amodiaquine, lumefantrine, and piperaquine.^27^ Polymorphisms in the ferrodoxine (*fd*), apicoplast ribosomal protein S10 (*arps10*), multidrug resistance protein 2 (*mdr2*) and *crt* genes have been described as a genetic background on which *pfk13* mutations are likely to arise.^28^

No *pfk13* mutations, but a low prevalence of *pfcrt* and *pfmdr1* mutations were detected in Namibia from 2016 to 2017.^29^ More recent unpublished data from 2019-2020 has identified the *pfk13* P441L mutation among malaria parasites from agricultural workers in Zambezi region (unpublished data). As part of the multi-country Genomics of Malaria in the Elimination 8 (GenE8) initiative, malaria molecular surveillance was conducted in Angola, Eswatini, Namibia, South Africa and Zambia, using a targeted amplicon sequencing workflow. Here, we report on antimalarial resistance markers present in samples collected in 16 health facilities along Namibia’s northern border between April and September 2023.

## Materials and Methods

### Study site

The study took place in 5 high transmission regions (Omusati, Ohangwena, Kavango West, Kavango East, and Zambezi) in northern Namibia (Figure 1). These regions border Angola, Botswana, and Zambia. Transmission is highly seasonal, occurring between December to June, with cases peaking between February and April during the rainy season.^30^ *P*. *falciparum* accounts for over 90% of infections, while other less prevalent non-falciparum species, such as *Plasmodium ovale*, *Plasmodium vivax*, and *Plasmodium malariae*, are also present.^4,9,29^ The most common vector is *Anopheles arabiensis*.^1,5^

**Figure 1.**
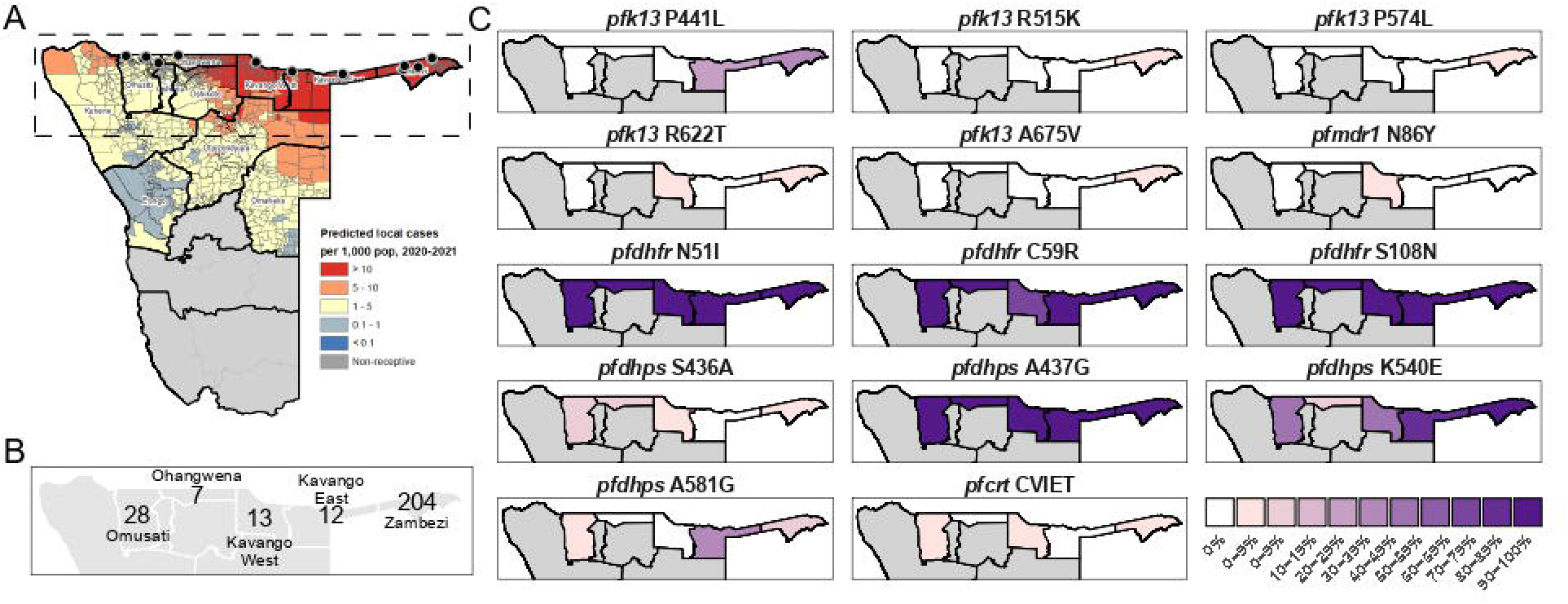
Proportion of samples carrying antimalarial drug resistance markers in Northern Namibia. **A**. Map of Namibia with risk zones stratified by predicted local cases per 1,000 population in 2020-2021. Data collection was conducted at health facilities indicated in black dots from April to September 2023. **B**. Number of sequenced samples per region. **C.** Proportion of samples carrying resistance markers of interest per region. Other mutations and source data can be found in the Supplementary Material.

### Study Design and Sampling

This was a cross-sectional study nested within the national surveillance system and conducted as part of the wider GenE8 project. The sampling strategy prioritized representativeness across geographies and transmission strata based on ecology and incidence of malaria, optimizing resource allocation to cover a significant proportion of the expected cases. The study targeted a minimum of 50 malaria-positive patients across 16 health facilities along Namibia’s northern border (Figure 1) in Omusati, Ohangwena, Kavango East, Kavango West and Zambezi regions. The target sample size (n=800) was intended to: a) estimate a 5% prevalence of a given molecular marker with a precision of 2.0% and 95% confidence, and b) achieve 90% power to detect a prevalence exceeding a value of 2% if the true prevalence is 4.3% using a one-sided binomial test with =0.05 and a design effect of 1.5. The achieved sample size of 264 provided a precision of 2.6% for a proportion of 5%. Sampling was conducted through passive case detection during part of a single transmission season (April–September 2023). Symptomatic patients presenting to participating health facilities were tested with a Pf/Pan RDT (MERISCREEN Malaria Pf/Pan Ag, Meril Diagnostics) for the detection of *P. falciparum* histidine-rich protein 2 (HRP2) and common *Plasmodium* lactate dehydrogenase (pLDH) according to national guidelines.^12^ All patients testing positive for malaria and aged over 2 years were invited to participate. Any participant who had taken malaria prophylaxis or treatment within the preceding 14 days was excluded from the study.

### Sample Collection

Written consent was obtained as described in Ethical Considerations prior to recruitment. Consenting participants were interviewed by trained study staff at the point of care in the health facility using pre-programmed survey tools on encrypted Android tablets that captured information on demographics, clinical symptoms, travel history, occupation and routine malaria interventions used by the participant or in their residence (including bed net usage, chemoprophylaxis and indoor residual spraying). Dried blood spots (DBS) collected from finger prick blood samples were uniquely labeled with barcodes to enable the linking of genomic and survey data, air dried, and stored at −20 in sealed bags with desiccants until laboratory processing. All participants were treated with the first-line treatment for uncomplicated malaria (artemether-lumefantrine) in line with national guidelines.^12^

### DNA extraction, qPCR and Multiplexed Amplicon Sequencing

All laboratory procedures were conducted at the National Institute for Communicable Diseases in Johannesburg, South Africa. Only DBS samples that could be linked to their epidemiological data were added to the sequencing workflow. DNA was extracted from a 6-mm hole-punch from DBS using the Tween20–Chelex protocol. 150 uL 10% Chelex 100 Resin in water was used for the final extraction step and the supernatant was transferred to DNA storage tubes.^31^ Parasite density was estimated by quantitative PCR targeting the *varATS* gene.^32^ Samples with < 5 parasite/μL were excluded from further processing. Sequencing libraries were prepared using an assay based on multiplexed amplicons for drug, diagnostic, diversity, and differentiation using high throughput targeted resequencing, MAD^4^HatTeR.^33^ Briefly, a multiplexed PCR using primer pools D1.1, R1.1 and R1.2 was performed with 15 or 20 cycles (for samples with parasitemia ≥10 and <10 parasite/μL, respectively). The multiplexed PCR was followed by bead cleaning, digestion and an indexing PCR following the CleanPlex Amplicon Sequencing protocol and using Paragon Genomics reagents. Indexing PCR was performed for 15 cycles using dual unique indexes. Library quality was checked on a random subset of samples using capillary electrophoresis (Tapestation 4200, Agilent Technologies, using D1000 ScreenTapes and reagents). Samples were pooled with volumes inversely proportional to the sample parasitemia, and pools of samples were combined to run on a NextSeq 2000 instrument using 150 paired-end reads, with at most 384 libraries per run using a P2 reagent kit.

### Bioinformatics Data Processing

The resulting demultiplexed fastq files were processed by a custom bioinformatic pipeline to infer alleles using version v0.2.1 of the pipeline, available on github.com/EPPIcenter/mad4hatter, and further data cleaning was conducted in R (version 4.3.1).^33^ Briefly, reads were demultiplexed into each target, filtered (by quality, length, and to remove primer dimers), and amplicon sequences were inferred using DADA2. Resulting amplicon sequence variants were then aligned to the reference genome (*P*. *falciparum* 3D7 version 2020-09-01) to filter out unspecific amplification. To improve precision and specificity we used the following DADA2 parameters: OMEGA_A=1e-120, pool=pseudo. To select priors for pseudo-pooling in the second round of inference, PSEUDO_ABUNDANCE=100, and PSEUDO_PREVALENCE=1000 were used which effectively constrained it to alleles observed with more than 100 reads in any sample. Target sequences in the *pfk13*, *pfcrt*, *pfmdr1*, *pfmdr2*, *pfdhps*, *pfdhfr*, *pfcoronin*, *arps10*, *pib7*, *fd*, *exo*, and PF3D7_1322700 genes were assessed. SNPs contained within the same amplicon are reported as microhaplotypes.

Samples with less than 10,000 reads, targets with ≤50 reads and SNPs with within-sample-allele-frequency ≤1% or ≤10 reads were excluded from the analysis. Non*-falciparum* targets were excluded from sequencing depth summaries. Otherwise, all resulting data were included, including samples with partial coverage of SNPs, resulting in varying total sample numbers for each SNP.

### Data Analysis

Further data analysis and visualizations were conducted in R (version 4.3.1). Farming and herding occupations were combined into a single category (agricultural workers), while all other occupations, excluding minors, students, and the unemployed, were grouped together (other workers). Additionally, to enable statistical analysis, individuals aged 25 years or older were grouped into a single category, and travel reports (within the 2 months prior to sample collection) were categorized as either international or domestic.

The proportion of samples carrying a mutation (i.e. polyclonal samples with mixed genotypes counted as a carrier of the mutation) was calculated for each health facility, region and overall, along with their 95% confidence intervals, calculated using the binom.test function in R. Multivariate logistic regression was performed using a generalized linear mixed-effects model to assess associations between the presence of markers of interest (as a binary outcome) and fixed effects including age, gender, residence status and travel history. The model incorporated a random intercept for each region. This analysis was restricted to variables with sufficient sample size for convergence. A Bayesian hierarchical logistic multivariate regression model with a random effect for health facility was fit to evaluate the geographical patterns of markers of interest.

Multilocus haplotypes were constructed for polymorphisms in *pfmdr1* (amino acids 86, 184 and 1246, targeted by 3 independent amplicons), and *pfdhpf*/*pfdhps* (amino acids in *pfdhps*: 436, 437, 540 and 581, and in *pfdhfr*: 51 and 59, targeted by 4 independent amplicons). Samples with a mixed genotype in 1 of the amplicons were considered to contain both haplotypes; samples with mixed genotypes in > 2 amplicons were considered undetermined.

The complexity of infection for each sample was calculated as the number of alleles corresponding to the 95^th^ percentile of the allele counts per locus within the sample. Population complexity of infection (COI) and 95% confidence intervals were obtained by Poisson regression. Allele frequencies were then estimated considering the COI of each sample by maximum likelihood estimation. One-dimensional likelihood ratio-based 95% confidence intervals were estimated for the minor allele.

Species-specific *ldh* targets in the panel were used to identify co-infections with non-*falciparum* species. The resulting sequences were used to remove unspecific amplification and to classify *P*. *ovale* reads into *P*. *ovale curtisi* or *P*. *ovale wallikeri*. Non-*falciparum* species were considered present if the combined reads from all *ldh* targets were > 100, with each species contributing > 1% of the total *ldh* targets reads.

### Ethical Considerations

Ethics approval was obtained from the Namibian Ministry of Health and Social Services Biomedical Research Ethics Committee (22/4/2/3) and the University of Namibia’s Research Ethics Committee (UNAM-DEC-MRS-008--24.08.2022). Written informed consent was obtained from all patients aged ≥18 years, as well as parents/guardians of children aged 2 to 11 years. Patients aged ≥12 to <18 years had to provide assent in addition to parent/guardian consent. The original signed consent forms were kept on file with a copy shared with the participants or parents/guardians. All personal identifiers were removed from survey data prior to secure storage on a password-protected cloud, while consent forms were kept in a restricted-access location.

## Results

The 16 selected health facilities reported a total of 1,246 malaria cases during the study period. Of 278 DBS collected, 263 with complete survey data were subjected to DNA extraction and quantification by qPCR. 11 samples with negative or missing RDT results (3 and 8, respectively) were included in the workflow (Table S1). A total of 264 samples with parasitemia above 5 parasites/μL were sequenced. 77.3% of sequenced samples came from Zambezi region and 56% from male participants (Table 1). Most study participants were Namibian residents (89%), with limited travel outside the habitual residence in the 2 months prior to enrolment in the study (7.3%). The largest proportion of participants fell within the 5–14 years age group (33%). Aside from minors, frequently reported occupation categories included students and the unemployed.

**Table 1.** Summary of demographic, geographic, and clinical data for individuals with sequenced samples, overall and by region.

| Characteristic | Overall | Region |  |  |  |  |
| --- | --- | --- | --- | --- | --- | --- |
|  |  | Omusati | Ohangwena | Kavango West | Kavango East | Zambezi |
| Health Facilities | 16 | 4 | 1 | 1 | 5 | 5 |
| Sequenced Samples | 264 | 28 (10.6%) | 7 (2.7%) | 13 (4.9%) | 12 (4.5%) | 204 (77.3%) |
| **Residence Country** |  |  |  |  |  |  |
| Angola | 25 (9.5%) | 18 (64%) | 4 (57%) | 3 (23%) | 0 (0%) | 0 (0%) |
| Namibia | 236 (89%) | 10 (36%) | 3 (43%) | 10 (77%) | 12 (100%) | 201(99%) |
| Zambia | 3 (1.1%) | 0 (0%) | 0 (0%) | 0 (0%) | 0 (0%) | 3 (1.5%) |
| **Age Group** |  |  |  |  |  |  |
| <5 | 24 (9.1%) | 4 (14%) | 0 (0%) | 2 (15%) | 0 (0%) | 18 (8.8%) |
| 5-14 | 88 (33%) | 5 (18%) | 3 (43%) | 4 (31%) | 5 (42%) | 71 (35%) |
| 15-24 | 59 (22%) | 5 (18%) | 1 (14%) | 1 (7.7%) | 3 (25%) | 49 (24%) |
| 25-39 | 55 (21%) | 7 (25%) | 1 (14%) | 5 (38%) | 2 (17%) | 40 (20%) |
| 40-59 | 26 (9.8%) | 3 (11%) | 2 (29%) | 1 (7.7%) | 0 (0%) | 20 (9.8%) |
| 60+ | 12 (4.5%) | 4 (14%) | 0 (0%) | 0 (0%) | 2 (17%) | 6 (2.9%) |
| **Gender** |  |  |  |  |  |  |
| Female | 116 (44%) | 16 (57%) | 0 (0%) | 6 (46%) | 2 (17%) | 92 (45%) |
| Male | 148 (56%) | 12 (43%) | 7 (100%) | 7 (54%) | 10 (83%) | 112 (55%) |
| **Travel History** |  |  |  |  |  |  |
| No Travel | 243 (93%) | 17 (63%) | 4 (57%) | 12 (100%) | 10 (83%) | 200 (98%) |
| Domestic | 19 (7.3%) | 10 (37%) | 3 (43%) | 0 (0%) | 2 (17%) | 4 (2.0%) |
| International | 2 | 1 | 0 | 1 | 0 | 0 |

### Sequencing performance and sequenced samples characteristics

Overall sequencing depth was high, with a median of 2,312,326 reads per sample (IQR: 561,791 - 4,451,199) and 4,051 reads per target (IQR: 1,163 – 12,919). Of the 264 samples, 253 and 144 had > 100 reads in 75% or 95% of the targets, respectively. Overall parasitemia was high, and successfully sequenced samples had higher parasitemia than unsuccessfully sequenced samples (median 65,652.4 parasite/μL, n = 254, and 8.7 parasite/μL, n = 10, respectively). More than half (53.5%) of the samples were polyclonal, with a population COI of 1.79 (95% CI: 1.68-1.90, N=254, Figure 2). COI was not correlated with either parasitemia or sequencing depth (*p* > 0.1) but was significantly lower in Kavango West and Omusati than Zambezi (*p* = 0.03 and 5.4×10^-5^, respectively). Non-*falciparum* species *P*. *ovale wallikeri*, *P*. *ovale curtisi* and *P. malariae* were detected in 5.68% of the samples (Table S2). *P*. *vivax* was not detected in any samples.

**Figure 2.**
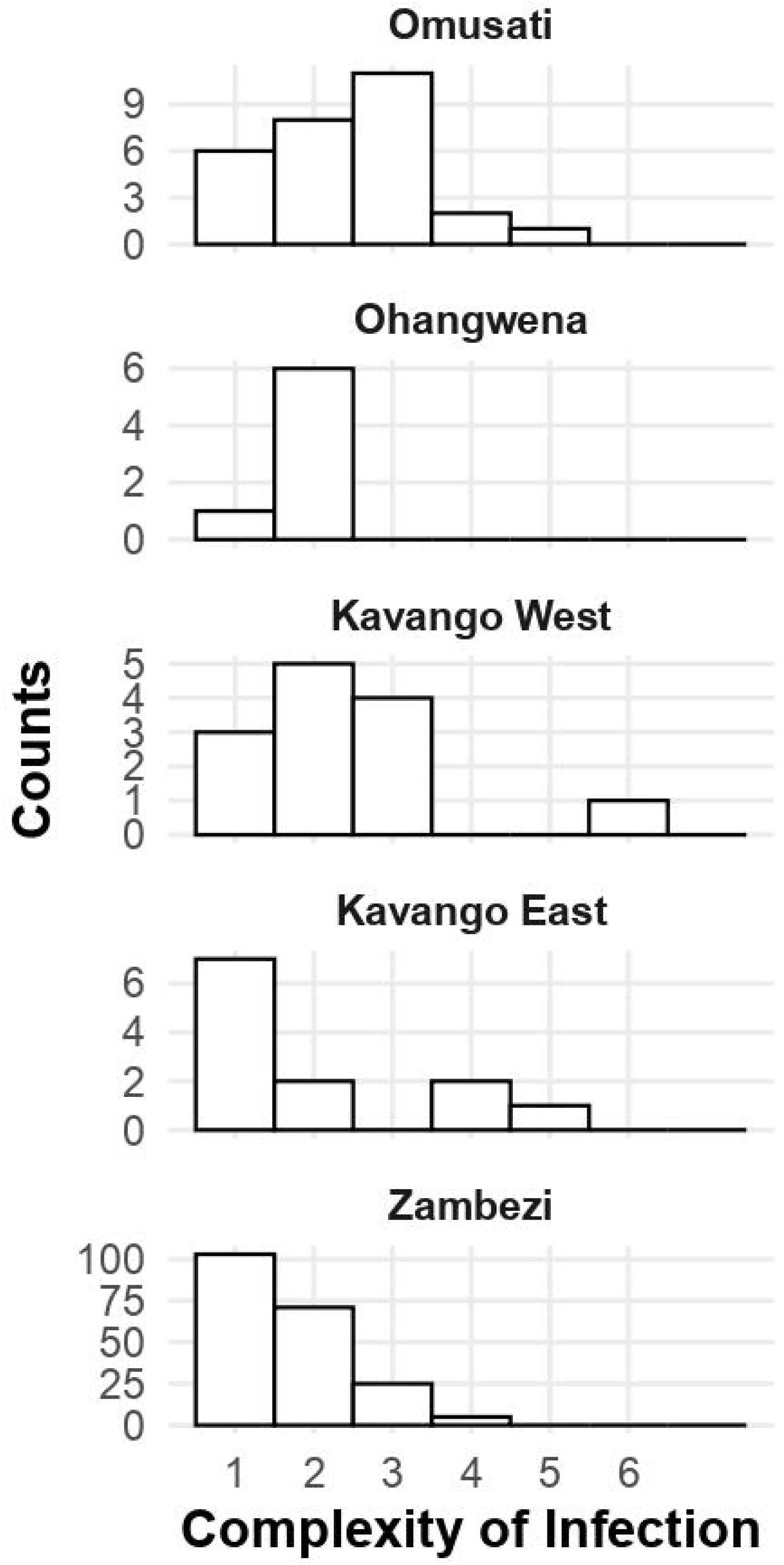
Distribution of complexity of infection (COI) for samples in each region.

### Antimalarial drug resistance molecular markers

Variants were identified in 7 genes: *arps10, pfcrt, pfdhfr, pfdhps, pfk13, pfmdr1,* and *pfmdr2*. No variants were detected in *PF3D7_13227700, coronin, exo, fd,* and *pib7* (Table S3).

Several mutations were detected in the *pfk13* gene, including candidate and validated markers of artemisinin partial resistance (Figure 1, Table S3-4). The candidate P441L marker was the most abundant mutation detected. Approximately a third of the samples carried this mutation (85 of 258 samples, 32.9% - 95% CI: 27.2-39.0%), with 4 samples found in Kavango East (36.4%, 95% CI: 10.9-69.2%), and 81 in Zambezi (40.1%, 95% CI: 33.3 - 47.2%). Health facilities within Zambezi had similar proportions of samples carrying the mutation (Table S5). 45.9% of the infections carrying P441L had a mixed genotype (Table S6), and those without a mixed genotype were not clonal. Males (56% of the total sample) were less likely to carry the marker, with an odds ratio of 0.54 (95% CI: 0.39-0.95, Table 2), and the mutation showed a geographical trend, increasing west to east (Spearman’s rank correlation: 0.77, *p* = 0.002, for the relationship between longitude and average rank of random effect per health facility). There was no evidence of an association between P441L and age, residency, or travel history. Two validated markers of artemisinin partial-resistance (P574L and A675V), as well as the candidate marker R515K, were detected in Zambezi region at low levels (Figure 1, Tables S4-6). Additional *pfk13* mutations observed in > 1% of the samples were a synonymous mutation, A504A (1.6%, CI: 0.4-4.1%), and an uncharacterized mutation, R622T (3.9%, CI: 1.9-7%).

**Table 2.** Multivariate logistic regression results for relevant demographics and drug resistance markers of interest.

| Characteristic | *pfk13* P441L<br>(85<sup>a</sup>/247<sup>b</sup>) |  | *pfdhps* K540E<br>(211<sup>a</sup>/249<sup>b</sup>) |  |
| --- | --- | --- | --- | --- |
|  | OR (95% CI) | *p* value | OR (95% CI) | *p* value |
| **Residence** |  | 0.2 |  | 0.6 |
| Namibian resident | Ref. |  | Ref. |  |
| Not a Namibian resident | 0.11 (0.0 - 2.74) |  | 0.65 (0.13 - 3.40) |  |
| **Age** |  | >0.9 |  | 0.2 |
| <5 | Ref. |  | Ref. |  |
| 5-14 | 1.37 (0.47 - 3.99) |  | 2.38 (0.72 - 7.80) |  |
| 15-24 | 1.21 (0.39 - 3.76) |  | 3.93 (0.98 - 15.8) |  |
| 25+ | 1.14 (0.38 - 3.36) |  | 2.91 (0.86 - 9.79) |  |
| **Gender** |  | **0.034** |  | 0.7 |
| Female | Ref. |  | Ref. |  |
| Male | 0.54 (0.30 - 0.95) |  | 1.18 (0.53 - 2.63) |  |
| **Travel history** |  | 0.5 |  | 0.13 |
| No travel | Ref. |  | Ref. |  |
| Domestic | 4.86 (0.36 – 66.1) |  | 0.67 (0.11 - 4.02) |  |
| International | 1.41 (0.10 – 20.3) |  | 0.17 (0.03 – 0.96) |  |

<sup>a</sup>: number of samples carrying the marker. <sup>b</sup>: total number of samples.

*Pfmdr1* N86Y, G182G and D1246Y were observed in < 2% of the samples (Figure 1, Tables S4-6). The *Pfmdr1* N86 was present in all samples and only one sample of mixed genotype carried the N86Y mutation. *Pfmdr1* Y184F (53.7%) and *pfmdr2* I492V (52.5%) were present in just over half of the samples and did not show a significant association with longitude (*p* = 0.88 and 0.39, respectively). The *pfmdr1* NYD (N86/Y184/D1246) haplotype was carried by the majority of samples (67%), followed by NFD (N86/184F/D1246, 50.6%, Table S7). The *pfcrt* 72-76 CVIET microhaplotype associated with chloroquine resistance was observed in 1.1% of all samples. Over 80% of samples carried the pyrimethamine resistance (*pfdhfr* N51I, C59R, and S108N), and sulphadoxine resistance (*pfdhps* A437G and K540E) markers. The quintuple mutant haplotype containing all those mutations was found in 67% of the samples, while the sextuple mutant haplotype containing, in addition, *pfdhps* A581G, was found in 8.7% of the samples (Table S7). As a single polymorphism, *pfdhps* A581G was found in 10.8% of the samples and, in all cases, occurred alongside *pfdhps* K540E within the same microhaplotype. 7.2% of the samples had undetermined *pfdhps*/*pfdhfr* haplotypes. *pfdhps* S436A was present in 3.4% of the samples, mostly in samples with a *pfdhps* K540 genotype. There were no significant associations of *pfdhps* K540E with any demographics, although its proportion was higher in eastern longitudes (Spearman’s rank correlation: 0.56, *p* = 0.042). Observed allele frequencies for all individual markers were lower than the proportion of samples carrying the allele (Table S8).

## Discussion

This manuscript reports the first published data using deep sequencing to characterize antimalarial drug resistance markers in Namibia, a southern African country with low and highly geographically concentrated transmission. The key findings include high levels of markers for artemisinin partial resistance, mutations associated with resistance to sulphadoxine and pyrimethamine, and the *Pfmdr1* N86 genotype, which is potentially selected by lumefantrine use. A candidate marker of artemisinin partial resistance, the *Pfk13* P441L mutation, was found in a high proportion of the samples, mainly in the Zambezi region (40.1%). This region is in the vicinity of 4 other countries: Angola, Botswana, Zambia and Zimbabwe. Approximately half of the infections were polyclonal, with the highest COI found in the lowest transmission region, Omusati. The genomic assay also detected a low proportion of co-infections with *P*. *malariae*, *P*. *ovale curtisi* and *P*. *ovale wallikeri*, but no *P*. *vivax*.

The study identified both ‘candidate’ and ‘validated’ *Pfk13* markers for artemisinin partial resistance in Namibia. The candidate marker *Pfk13* P441L mutation first found in the Greater Mekong Subregion, has been reported in western Uganda (up to 23% in one site), and at very low prevalence in Central Africa.^22,34^ This marker has been associated with delayed clearance in the Greater Mekong Subregion; however, in a study with very low sample sizes from that region, it showed no increased ring survival ex vivo.^35,36^ Its clinical efficacy and phenotypical effects in African parasites remains unassessed.^13^ Other low-level markers of artemisinin partial resistance found in this study have clinical and/or laboratory evidence in Africa; these include the rare markers P574L and R515K, and A675V, which has shown a notable increase in Uganda.^21,22,37^ Notably, the R622T mutation, observed in 3.9% of the samples, occurs at the same amino acid position as the validated marker R622I causing delayed clearance in the Horn of Africa, suggesting a potential similar phenotype for R622T.^24^

There was a distinct variation in the distribution of mutations between regions in northern Namibia. While the Zambezi region had a higher proportion of samples with any *Pfk13* mutation, there were limited samples from other regions to generate reliable estimates. Our results did not find a significant association between residence in Namibia or travel history and P441L carriage, and data collection occurred late in the transmission season. Nevertheless, most samples corresponded to Namibian residents and participants who did not report travel (84 and 82 out of 85 P441L carriers, respectively) and these infections were not clonal, suggesting that multiple parasites carrying the mutation are sustained by local transmission. High parasite connectivity between Namibia, Angola and Zambia, driven by human mobility, suggests that this mutation is likely to be found in neighboring countries.^6,38,39^ The only significant association observed with P441L carriage was a lower likelihood among males. These results could be attributed to potential misreporting of variables such as travel history, or due to correlations with other characteristics that lacked sufficient sample sizes for regression analysis. Sample sizes were not substantial enough to identify associations for other *Pfk13* mutations, although all participants carrying them were residents and reported no travel. None of the markers described in this study were identified in a survey conducted in 2017; however, unpublished data from 2020 identified the P441L marker in cross-border and resident agricultural workers.

The interplay between transmission intensity, host immunity and pharmacokinetics, drug pressure and parasite physiology play a critical role in shaping the emergence and propagation of drug resistance.^40^ Historically, chloroquine resistance first developed in areas of low malaria transmission, and antifolate resistance has been observed to spread more swiftly in low-transmission regions compared to those with high transmission.^40^ Regions that have experienced the emergence of artemisinin partial resistance are either low transmission areas or areas that experienced an upsurge in cases.^22,41^ In Namibia, a low transmission setting, multiple factors may have contributed to the selection and increase of artemisinin partial resistance markers to the levels described in this study. Recent epidemics may have driven selection by exposing non-immune populations to malaria, increasing treatment-seeking behavior and, consequently, drug pressure.^13,22^ Modeling analyses have suggested that the risk of selecting for drug resistance parasites is high in Namibia given its low incidence and high artemether-lumefantrine usage. ^42^ Lower population immunity due to reduced exposure could also have limited the hosts’ ability to clear drug-resistant parasites despite potential fitness costs.^13^ Additionally, low transmission is typically associated with lower polyclonality (85.3% of the infections in Zambezi had a COI ≤ 2), reducing within-host parasite competition.^13,26^ Despite limited sample sizes, the COI was higher in Omusati, a region with very low transmission, likely reflecting importation from high transmission areas and underscoring the potential impact of imported cases on local parasite diversity. Other potential contributing factors include increased presumptive treatment due to false RDT positives and the use of artemisinin-based antimalarials for targeted mass drug administration in Namibia and Zambia, leading to unnecessary drug pressure.^43–45^ Retrospective studies are necessary to describe the emergence dynamics of these markers and to shed light on the contribution of some of these factors at the time the markers propagated.

While artemisinin partial resistance may not lead to ACT failures due to the action of the partner drugs, it threatens the efficacy of intravenous artesunate monotherapies used to treat severe malaria.^13^ Reduced efficacy of the artemisinin component also puts additional pressure on the partner drug and could lead to ACT failure in the presence of resistance to the partner drug.^46^ Ensuring the use of efficacious partner drugs is therefore critical. No evidence of full lumefantrine resistance has been documented in Africa, but the high prevalence of *pfmdr1* N86 and *pfcrt* K76 genotypes may be linked to a modest reduction in lumefantrine susceptibility.^15,47^ Conversely, the *pfcrt* 72-76 CVIET microhaplotype, along with the *pfmdr1* N86Y mutation, are associated with resistance to chloroquine and, to a lesser extent, amodiaquine.^48^ These differences in susceptibility may be behind the differential selection of the *pfmdr1* NFD (N86/184F/D1246) and YYY (86Y/Y184/1246Y) haplotypes by artemether-lumefantrine and artesunate-amodiaquine use, respectively.^27^ Our data shows that the *pfmdr1* N86 (mostly in NFD and NYD haplotypes) and *pfcrt* K76 are ubiquitous. These patterns are likely shaped by the discontinuation of chloroquine use in Namibia after resistance was detected in 1984, and the heavy reliance on artemether-lumefantrine.^11^ No mutations in *pfcrt* associated with piperaquine resistance were detected.

Sulphadoxine-pyrimethamine resistance is driven by accumulated mutations in *pfdhps* and *pfdhfr*.^27,49^ The quintuple mutant haplotype is associated with treatment resistance, and the sextuple mutant haplotype confers even higher levels of resistance.^50^ The quintuple mutant haplotype was found in a majority of the samples, and its hallmark mutation, *pfdhps* K540E, was found at very high levels. The sextuple mutant haplotype containing *pfdhps* A581G was observed in approximately a tenth of the samples. Other sextuple mutations, *pfdhps* A613S/T and *pfdhfr* I164L, were absent. These results are notable, as sulphadoxine-pyrimethamine use was discontinued in Namibia following the detection of resistance in 2004.^11^ However, its use for chemoprevention in neighboring Angola, Zambia and Zimbabwe may sustain the selection of these markers in this highly interconnected region.^6^ Similar to sulphadoxine-pyrimethamine, cotrimoxazole, widely used for prophylaxis in human immunodeficiency virus-infected individuals, targets the folate biosynthesis pathway in *P*. *falciparum*. Thus, the use of cotrimoxazole may contribute to the high levels of sulphadoxine-pyrimethamine resistance markers found in this study.^51^ Overall, these data indicate that sulphadoxine-pyrimethamine should not be recommended for treatment.

This study had several limitations. Sample collection commenced mid-way through the malaria season due to administrative delays, leading to a lower sample size. This limited statistical inference and use of regression models for risk factors, and precluded further investigations around importation and transmission patterns early in the season. Sample sizes were higher in the Zambezi region, leading to a higher degree of confidence in the representativeness of findings within the region. Samples were collected exclusively from symptomatic patients at health facilities, potentially underrepresenting parasite populations in asymptomatic infections, recent travelers, and users of traditional and other medicines. Short amplicon sequencing presents challenges in phasing SNPs to construct multilocus haplotypes; to mitigate biases from ambiguous polyclonal samples, our analysis predominantly focused on individual SNPs. Finally, the finding of non-*falciparum* species in Namibia requires further validation by other methods, given the assay’s sensitivity and specificity are not well-characterized for this particular application.

Achieving malaria elimination in Namibia requires continuous, effective interventions, yet the threat of antimalarial drug resistance remains significant. This study highlights a high prevalence of resistance markers, underscoring the urgent need for further research to evaluate their phenotypic and clinical impacts, including the efficacy of ACTs. The findings also emphasize the importance of molecular surveillance in detecting emerging threats, advocating for routine, geographically representative monitoring of both known, novel and uncharacterized mutations. To sustain progress, future efforts should focus on strengthening health systems, improving data collection, and fostering community engagement, ensuring the long-term success of malaria control and eventual elimination.

## Data Availability

All data produced in the present study are available upon reasonable request to the authors

## Acknowledgments

We thank the participants, nurses, field supervisors and data managers that made this study possible. We also thank members of the Malaria Elimination Initiative, the EPPIcenter and Johns Hopkins University’s IDDynamics group - especially Bryan Greenhouse, Inna Gerlovina and Alfred Hubbard - for fruitful technical discussions.

## Co-Author Contact Information

Lydia Eloff. Department of Biochemistry, Microbiology, and Biotechnology, School of Science, University of Namibia, P/Bag 13301, Windhoek, Namibia, Andrés Aranda-Díaz. EPPIcenter Research Program, Department of Medicine, University of California, San Francisco, United States of America

Isobel Routledge. Malaria Elimination Initiative, Global Health Group, University of California, San Francisco, United States of America

Jennifer Smith. Malaria Elimination Initiative, Global Health Group, University of California, San Francisco, United States of America

Mukosha Chisenga. SADC Malaria Elimination Eight Secretariat, Windhoek, Namibia

Brighton Mangena. SADC Malaria Elimination Eight Secretariat, Windhoek, Namibia

Amy Wesolowski. Department of Epidemiology, Johns Hopkins University Bloomberg School of Public Health, Baltimore, Maryland, United States of America. Chadwick Sikala. SADC Malaria Elimination Eight Secretariat, Windhoek, Namibia

John Chimumbwa. SADC Malaria Elimination Eight Secretariat, Windhoek, Namibia

Stark Katokele. National Vector-Borne Disease Control Program, Ministry of Health and Social Services, Harvey Street, Private Bag 13198, Windhoek, Namibia

Petrina Uusiku. National Vector-Borne Disease Control Program, Ministry of Health and Social Services, Harvey Street, Private Bag 13198, Windhoek, Namibia, Jaishree Raman. Laboratory for Antimalarial Resistance Monitoring and Malaria Operational Research (ARMMOR), Centre of Emerging Zoonotic and Parasitic Diseases, National Institute for Communicable Diseases; Wits Research Institute for Malaria, University of Witwatersrand; UP Institute for Sustainable Malaria Control, University of Pretoria; Johannesburg, South Africa

Davis R. Mumbengegwi. Malaria Operational Research Program, Centre for Research Services, University of Namibia, P/Bag 13301, Windhoek, Namibia

