## Supplemental tables1-8 for "High Prevalence of Molecular Markers Associated with Artemisinin, Sulphadoxine and Pyrimethamine Resistance in Northern Namibia"

| | |
|---|---|
| Table S1 | Workflow of processed and sequenced samples |
| Table S2 | Distribution of *Plasmodium* species in *P . falciparum* mono- and co-infections overall anc |
| Table S3 | Summary of observed variation in SNPs of interest within the full dataset. |
| Table S4 | Overall and region-level proportion of infections carrying a mutation of interest |
| Table S5 | Proportion of infections carrying a mutation of interest in health facilities in Zambezi regi |
| Table S6 | Proportion of sample genotypes (reference, alternate and mixed) by region and overall |
| Table S7 | Proportion of samples carrying a haplotype of interest |
| Table S8 | Allele frequencies for minor alleles, adjusted by the complexity of infection of each samp |

**Table S1. Workflow of processed and sequenced samples**

| RDT result | DNA extraction | Parasitemia > 5 parasite/uL | Succesfully sequenced* |
|---|---|---|---|
| **Positive** | 263 | 253 | 253 |
| **Negative**** | 3 | 3 | 3 |
| **Missing**** | 8 | 8 | 8 |
| **Total** | 274 | 264 | 264 |

*: Samples with >10,000 reads. Samples with partial coverage of SNPs are still considered succesfully sequenced

**: Samples exceptionally included in the workflow

**Table S2. Distribution of *Plasmodium* species in *P . falciparum* mono- and co-infections overall and by region**

| Species | Overall (N = 264) | Kavango East (N = 12) | Kavango West (N = 13) | Ohangwena (N = 7) | Omusati (N = 28) | Zambezi (N = 204) |
|---|---|---|---|---|---|---|
| **Pf** | 249 (94.32%) | 12 (100%) | 12 (92.31%) | 5 (71.43%) | 25 (89.29%) | 195 (95.59%) |
| **Pf +Pm** | 6 (2.27%) | 0 (0%) | 1 (7.69%) | 0 (0%) | 2 (7.14%) | 3 (1.47%) |
| **Pf +Pm+Pow** | 2 (0.76%) | 0 (0%) | 0 (0%) | 0 (0%) | 1 (3.57%) | 1 (0.49%) |
| **Pf +Poc** | 1 (0.38%) | 0 (0%) | 0 (0%) | 0 (0%) | 0 (0%) | 1 (0.49%) |
| **Pf +Pow** | 6 (2.27%) | 0 (0%) | 0 (0%) | 2 (28.57%) | 0 (0%) | 4 (1.96%) |

**Table S3. Summary of observed variation in SNPs of interest within the full dataset.**

Polyclonal samples with mixed genotypes count as carrying the mutation. SNPs where no samples with a mutation were observed are reported as “No variant”; SNPs with variation within the dataset are reported as “Variant”. Mutations observed in at most 1 or 2 samples are denoted with * and **, respectively.

| Gene | Variants | No Variants |
|---|---|---|
| arps10 | V127L* | D128 |
| crt | M74I; N75E; K76T | F48; F52; C72; V73; T93; H97; C101; F145; I218; A220; M343; C350; G353; I356 |
| dhfr | N51I; C59R; S108N | A16; I164; T185 |
| dhps | S436A; S436F*; A437G; K540E; A581G | I431; A613 |
| k13 | P441L; K479I**; A504A; R515K**; P574L; A578S*; E596V*; R622T; P667A**; P667S*; A675V; A675A* | G436; I437; F442; F446; G449; D452; V454; N458; C469; M476; K480; A481; F483; S485; Y493; V494; V510; V520; S522; P527; N537; G538; R539; I540; I543; G544; Y546; P553; V555; R561; V568; R575; C580; V581; D584; R597; L598; N599; E605; K607; Q613; L631; D641; F656; R659; Q661; F662; F673; G690 |
| mdr1 | N86Y*; G182G; Y184F; D1246Y | W186; T371; S1034; N1042 |
| mdr2 | I492V | D463; T484; I515 |
| PF3D7_1322 | | T236 |
| coronin | | G50; R100; E107 |
| exo | | E415 |
| fd | | D193 |
| pib7 | | C1484 |

**Table S4. Overall and region-level proportion of infections carrying a mutation of interest**

This table only includes SNPs observed at > 1% nationwide (for pfk13, candidate (*) and validated (**) markers are included regardless) in all regions

| | | | | Overall | | Omusati | | Ohangwena | | Kavango West | | Kavango East | | Zambezi | |
|---|---|---|---|---|---|---|---|---|---|---|---|---|---|---|---|
| **Gene** | **CodonID** | **Ref** | **Alt** | **n/N** | **% (95% CI)** | **n/N** | **% (95% CI)** | **n/N** | **% (95% CI)** | **n/N** | **% (95% CI)** | **n/N** | **% (95% CI)** | **n/N** | **% (95% CI)** |
| crt | 72-76 | CVMNK | CVIET | 3/262 | 1.1% (0.2% - 3.3%) | 1/28 | 3.6% (0.1% - 18.3%) | 0/7 | 0% (0% - 41%) | 1/13 | 7.7% (0.2% - 36%) | 0/12 | 0% (0% - 26.5%) | 1/202 | 0.5% (0% - 2.7%) |
| dhfr | 51 | N | I | 253/257 | 98.4% (96.1% - 99.6%) | 27/27 | 100% (87.2% - 100%) | 6/6 | 100% (54.1% - 100%) | 13/13 | 100% (75.3% - 100%) | 11/11 | 100% (71.5% - 100%) | 196/200 | 98% (95% - 99.5%) |
| dhfr | 59 | C | R | 252/257 | 98.1% (95.5% - 99.4%) | 26/27 | 96.3% (81% - 99.9%) | 6/6 | 100% (54.1% - 100%) | 10/13 | 76.9% (46.2% - 95%) | 11/11 | 100% (71.5% - 100%) | 199/200 | 99.5% (97.2% - 100%) |
| dhfr | 108 | S | N | 261/261 | 100% (98.6% - 100%) | 27/27 | 100% (87.2% - 100%) | 7/7 | 100% (59% - 100%) | 13/13 | 100% (75.3% - 100%) | 12/12 | 100% (73.5% - 100%) | 202/202 | 100% (98.2% - 100%) |
| dhps | 436 | S | A | 10/261 | 3.8% (1.9% - 6.9%) | 4/27 | 14.8% (4.2% - 33.7%) | 1/7 | 14.3% (0.4% - 57.9%) | 1/13 | 7.7% (0.2% - 36%) | 0/11 | 0% (0% - 28.5%) | 4/203 | 2% (0.5% - 5%) |
| dhps | 437 | A | G | 262/262 | 100% (98.6% - 100%) | 28/28 | 100% (87.7% - 100%) | 7/7 | 100% (59% - 100%) | 13/13 | 100% (75.3% - 100%) | 11/11 | 100% (71.5% - 100%) | 203/203 | 100% (98.2% - 100%) |
| dhps | 540 | K | E | 217/261 | 83.1% (78% - 87.5%) | 16/28 | 57.1% (37.2% - 75.5%) | 1/6 | 16.7% (0.4% - 64.1%) | 7/13 | 53.8% (25.1% - 80.8%) | 9/11 | 81.8% (48.2% - 97.7%) | 184/203 | 90.6% (85.8% - 94.3%) |
| dhps | 581 | A | G | 28/263 | 10.6% (7.2% - 15%) | 1/28 | 3.6% (0.1% - 18.3%) | 0/7 | 0% (0% - 41%) | 0/13 | 0% (0% - 24.7%) | 5/12 | 41.7% (15.2% - 72.3%) | 22/203 | 10.8% (6.9% - 15.9%) |
| k13 | 441 | P | L* | 85/258 | 32.9% (27.2% - 39%) | 0/27 | 0% (0% - 12.8%) | 0/6 | 0% (0% - 45.9%) | 0/12 | 0% (0% - 26.5%) | 4/11 | 36.4% (10.9% - 69.2%) | 81/202 | 40.1% (33.3% - 47.2%) |
| k13 | 504 | A | A | 4/248 | 1.6% (0.4% - 4.1%) | 0/27 | 0% (0% - 12.8%) | 0/7 | 0% (0% - 41%) | 0/12 | 0% (0% - 26.5%) | 0/11 | 0% (0% - 28.5%) | 4/191 | 2.1% (0.6% - 5.3%) |
| k13 | 515 | R | K* | 2/248 | 0.8% (0.1% - 2.9%) | 0/27 | 0% (0% - 12.8%) | 0/7 | 0% (0% - 41%) | 0/12 | 0% (0% - 26.5%) | 0/11 | 0% (0% - 28.5%) | 2/191 | 1% (0.1% - 3.7%) |
| k13 | 574 | P | L** | 3/263 | 1.1% (0.2% - 3.3%) | 0/28 | 0% (0% - 12.3%) | 0/7 | 0% (0% - 41%) | 0/13 | 0% (0% - 24.7%) | 0/12 | 0% (0% - 26.5%) | 3/203 | 1.5% (0.3% - 4.3%) |
| k13 | 622 | R | T | 10/258 | 3.9% (1.9% - 7%) | 0/27 | 0% (0% - 12.8%) | 0/6 | 0% (0% - 45.9%) | 1/13 | 7.7% (0.2% - 36%) | 0/12 | 0% (0% - 26.5%) | 9/200 | 4.5% (2.1% - 8.4%) |
| k13 | 675 | A | V** | 3/257 | 1.2% (0.2% - 3.4%) | 0/27 | 0% (0% - 12.8%) | 0/6 | 0% (0% - 45.9%) | 0/13 | 0% (0% - 24.7%) | 0/12 | 0% (0% - 26.5%) | 3/199 | 1.5% (0.3% - 4.3%) |
| mdr1 | 86 | N | Y | 1/255 | 0.4% (0% - 2.2%) | 0/27 | 0% (0% - 12.8%) | 0/5 | 0% (0% - 52.2%) | 1/13 | 7.7% (0.2% - 36%) | 0/10 | 0% (0% - 30.8%) | 0/200 | 0% (0% - 1.8%) |
| mdr1 | 182 | G | G | 4/259 | 1.5% (0.4% - 3.9%) | 0/27 | 0% (0% - 12.8%) | 0/7 | 0% (0% - 41%) | 0/13 | 0% (0% - 24.7%) | 0/11 | 0% (0% - 28.5%) | 4/201 | 2% (0.5% - 5%) |
| mdr1 | 184 | Y | F | 139/259 | 53.7% (47.4% - 59.9%) | 15/27 | 55.6% (35.3% - 74.5%) | 2/7 | 28.6% (3.7% - 71%) | 5/13 | 38.5% (13.9% - 68.4%) | 6/11 | 54.5% (23.4% - 83.3%) | 111/201 | 55.2% (48.1% - 62.2%) |
| mdr1 | 1246 | D | Y | 5/260 | 1.9% (0.6% - 4.4%) | 0/27 | 0% (0% - 12.8%) | 0/7 | 0% (0% - 41%) | 1/13 | 7.7% (0.2% - 36%) | 0/12 | 0% (0% - 26.5%) | 4/201 | 2% (0.5% - 5%) |
| mdr2 | 492 | I | V | 136/259 | 52.5% (46.2% - 58.7%) | 16/27 | 59.3% (38.8% - 77.6%) | 2/6 | 33.3% (4.3% - 77.7%) | 8/13 | 61.5% (31.6% - 86.1%) | 9/12 | 75% (42.8% - 94.5%) | 101/201 | 50.2% (43.1% - 57.4%) |

**Table S5. Proportion of infections carrying a mutation of interest in health facilities in Zambezi region**

This table only includes SNPs observed at > 1% nationwide (for pfk13, candidate (*) and validated (**) markers are included regardless) in all regions

| | | | | Choi Clinic | | Katima Mulilo Hospital | | Ngweze Clinic | | Sesheke Clinic | | Sibbinda Clinic | |
|---|---|---|---|---|---|---|---|---|---|---|---|---|---|
| **Gene** | **CodonID** | **Ref** | **Alt** | **n/N** | **% (95% CI)** | **n/N** | **% (95% CI)** | **n/N** | **% (95% CI)** | **n/N** | **% (95% CI)** | **n/N** | **% (95% CI)** |
| crt | 72-76 | CVMNK | CVIET | 0/125 | 0% (0% - 2.9%) | 0/14 | 0% (0% - 23.2%) | 0/8 | 0% (0% - 36.9%) | 0/31 | 0% (0% - 11.2%) | 1/24 | 4.2% (0.1% - 21.1%) |
| dhfr | 51 | N | I | 120/123 | 97.6% (93% - 99.5%) | 14/14 | 100% (76.8% - 100%) | 8/8 | 100% (63.1% - 100%) | 31/31 | 100% (88.8% - 100%) | 23/24 | 95.8% (78.9% - 99.9%) |
| dhfr | 59 | C | R | 123/123 | 100% (97% - 100%) | 14/14 | 100% (76.8% - 100%) | 8/8 | 100% (63.1% - 100%) | 31/31 | 100% (88.8% - 100%) | 23/24 | 95.8% (78.9% - 99.9%) |
| dhfr | 108 | S | N | 125/125 | 100% (97.1% - 100%) | 14/14 | 100% (76.8% - 100%) | 8/8 | 100% (63.1% - 100%) | 31/31 | 100% (88.8% - 100%) | 24/24 | 100% (85.8% - 100%) |
| dhps | 436 | S | A | 2/125 | 1.6% (0.2% - 5.7%) | 0/14 | 0% (0% - 23.2%) | 1/9 | 11.1% (0.3% - 48.2%) | 0/31 | 0% (0% - 11.2%) | 1/24 | 4.2% (0.1% - 21.1%) |
| dhps | 437 | A | G | 125/125 | 100% (97.1% - 100%) | 14/14 | 100% (76.8% - 100%) | 9/9 | 100% (66.4% - 100%) | 31/31 | 100% (88.8% - 100%) | 24/24 | 100% (85.8% - 100%) |
| dhps | 540 | K | E | 113/125 | 90.4% (83.8% - 94.9%) | 11/14 | 78.6% (49.2% - 95.3%) | 8/9 | 88.9% (51.8% - 99.7%) | 30/31 | 96.8% (83.3% - 99.9%) | 22/24 | 91.7% (73% - 99%) |
| dhps | 581 | A | G | 15/125 | 12% (6.9% - 19%) | 1/14 | 7.1% (0.2% - 33.9%) | 1/9 | 11.1% (0.3% - 48.2%) | 3/31 | 9.7% (2% - 25.8%) | 2/24 | 8.3% (1% - 27%) |
| k13 | 441 | P | L* | 49/125 | 39.2% (30.6% - 48.3%) | 4/14 | 28.6% (8.4% - 58.1%) | 7/8 | 87.5% (47.3% - 99.7%) | 11/31 | 35.5% (19.2% - 54.6%) | 10/24 | 41.7% (22.1% - 63.4%) |
| k13 | 504 | A | A | 3/118 | 2.5% (0.5% - 7.3%) | 0/14 | 0% (0% - 23.2%) | 0/8 | 0% (0% - 36.9%) | 1/29 | 3.4% (0.1% - 17.8%) | 0/22 | 0% (0% - 15.4%) |
| k13 | 515 | R | K* | 0/118 | 0% (0% - 3.1%) | 0/14 | 0% (0% - 23.2%) | 0/8 | 0% (0% - 36.9%) | 2/29 | 6.9% (0.8% - 22.8%) | 0/22 | 0% (0% - 15.4%) |
| k13 | 574 | P | L** | 3/126 | 2.4% (0.5% - 6.8%) | 0/14 | 0% (0% - 23.2%) | 0/8 | 0% (0% - 36.9%) | 0/31 | 0% (0% - 11.2%) | 0/24 | 0% (0% - 14.2%) |
| k13 | 622 | R | T | 3/123 | 2.4% (0.5% - 7%) | 0/14 | 0% (0% - 23.2%) | 1/8 | 12.5% (0.3% - 52.7%) | 2/31 | 6.5% (0.8% - 21.4%) | 3/24 | 12.5% (2.7% - 32.4%) |
| k13 | 675 | A | V** | 2/122 | 1.6% (0.2% - 5.8%) | 0/14 | 0% (0% - 23.2%) | 0/8 | 0% (0% - 36.9%) | 1/31 | 3.2% (0.1% - 16.7%) | 0/24 | 0% (0% - 14.2%) |
| mdr1 | 86 | N | Y | 0/123 | 0% (0% - 3%) | 0/14 | 0% (0% - 23.2%) | 0/8 | 0% (0% - 36.9%) | 0/31 | 0% (0% - 11.2%) | 0/24 | 0% (0% - 14.2%) |
| mdr1 | 182 | G | G | 3/124 | 2.4% (0.5% - 6.9%) | 0/14 | 0% (0% - 23.2%) | 0/8 | 0% (0% - 36.9%) | 1/31 | 3.2% (0.1% - 16.7%) | 0/24 | 0% (0% - 14.2%) |
| mdr1 | 184 | Y | F | 66/124 | 53.2% (44.1% - 62.2%) | 7/14 | 50% (23% - 77%) | 5/8 | 62.5% (24.5% - 91.5%) | 19/31 | 61.3% (42.2% - 78.2%) | 14/24 | 58.3% (36.6% - 77.9%) |
| mdr1 | 1246 | D | Y | 4/124 | 3.2% (0.9% - 8.1%) | 0/14 | 0% (0% - 23.2%) | 0/8 | 0% (0% - 36.9%) | 0/31 | 0% (0% - 11.2%) | 0/24 | 0% (0% - 14.2%) |
| mdr2 | 492 | I | V | 60/124 | 48.4% (39.3% - 57.5%) | 6/14 | 42.9% (17.7% - 71.1%) | 4/8 | 50% (15.7% - 84.3%) | 19/31 | 61.3% (42.2% - 78.2%) | 12/24 | 50% (29.1% - 70.9%) |

**Table S6. Proportion of sample genotypes (reference, alternate and mixed) by region and overall**

Artemisinin partial resistance markers: *: candidate, **: validated.

| | | | | Overall | | | Omusati | | | Ohangwena | | | Kavango West | | | Kavango East | | | Zambezi | | |
|---|---|---|---|---|---|---|---|---|---|---|---|---|---|---|---|---|---|---|---|---|---|
| Gene | CodonID | Ref | Alt | Ref | Alt | Mixed | Ref | Alt | Mixed | Ref | Alt | Mixed | Ref | Alt | Mixed | Ref | Alt | Mixed | Ref | Alt | Mixed |
| crt | 72-76 | CVMNK | CVIET | 259 (98.85%) | 0 (0%) | 3 (1.15%) | 27 (96.43%) | 0 (0%) | 1 (3.57%) | 7 (100%) | 0 (0%) | 0 (0%) | 12 (92.31%) | 0 (0%) | 1 (7.69%) | 12 (100%) | 0 (0%) | 0 (0%) | 201 (99.5%) | 0 (0%) | 1 (0.5%) |
| dhfr | 51 | N | I | 4 (1.56%) | 242 (94.16%) | 11 (4.28%) | 1 (3.7%) | 19 (70.37%) | 7 (25.93%) | 0 (0%) | 4 (66.67%) | 2 (33.33%) | 3 (23.08%) | 5 (38.46%) | 5 (38.46%) | 0 (0%) | 11 (100%) | 0 (0%) | 1 (0.5%) | 196 (98%) | 3 (1.5%) |
| dhfr | 59 | C | R | 5 (1.95%) | 235 (91.44%) | 17 (6.61%) | 0 (0%) | 26 (96.3%) | 1 (3.7%) | 0 (0%) | 7 (100%) | 0 (0%) | 0 (0%) | 13 (100%) | 0 (0%) | 0 (0%) | 12 (100%) | 0 (0%) | 0 (0%) | 202 (100%) | 0 (0%) |
| dhfr | 108 | S | N | 0 (0%) | 260 (99.62%) | 1 (0.38%) | 0 (0%) | 24 (88.89%) | 3 (11.11%) | 0 (0%) | 6 (100%) | 0 (0%) | 0 (0%) | 11 (84.62%) | 2 (15.38%) | 0 (0%) | 11 (100%) | 0 (0%) | 4 (2%) | 190 (95%) | 6 (3%) |
| dhps | 436 | S | A | 251 (95.8%) | 4 (1.53%) | 7 (2.67%) | 23 (82.14%) | 2 (7.14%) | 3 (10.71%) | 6 (85.71%) | 1 (14.29%) | 0 (0%) | 12 (92.31%) | 0 (0%) | 1 (7.69%) | 11 (100%) | 0 (0%) | 0 (0%) | 199 (98.03%) | 1 (0.49%) | 3 (1.48%) |
| dhps | 437 | A | G | 0 (0%) | 259 (98.85%) | 3 (1.15%) | 0 (0%) | 28 (100%) | 0 (0%) | 0 (0%) | 7 (100%) | 0 (0%) | 0 (0%) | 13 (100%) | 0 (0%) | 0 (0%) | 11 (100%) | 0 (0%) | 0 (0%) | 200 (98.52%) | 3 (1.48%) |
| dhps | 540 | K | E | 44 (16.86%) | 182 (69.73%) | 35 (13.41%) | 12 (42.86%) | 4 (14.29%) | 12 (42.86%) | 5 (83.33%) | 0 (0%) | 1 (16.67%) | 6 (46.15%) | 2 (15.38%) | 5 (38.46%) | 2 (18.18%) | 9 (81.82%) | 0 (0%) | 19 (9.36%) | 167 (82.27%) | 17 (8.37%) |
| dhps | 581 | A | G | 235 (89.35%) | 18 (6.84%) | 10 (3.8%) | 27 (96.43%) | 0 (0%) | 1 (3.57%) | 7 (100%) | 0 (0%) | 0 (0%) | 13 (100%) | 0 (0%) | 0 (0%) | 7 (58.33%) | 2 (16.67%) | 3 (25%) | 181 (89.16%) | 16 (7.88%) | 6 (2.96%) |
| k13 | 441 | P | L* | 173 (67.05%) | 46 (17.83%) | 39 (15.12%) | 27 (100%) | 0 (0%) | 0 (0%) | 6 (100%) | 0 (0%) | 0 (0%) | 12 (100%) | 0 (0%) | 0 (0%) | 7 (63.64%) | 2 (18.18%) | 2 (18.18%) | 121 (59.9%) | 44 (21.78%) | 37 (18.32%) |
| k13 | 504 | A | A | 244 (98.39%) | 3 (1.21%) | 1 (0.4%) | 27 (100%) | 0 (0%) | 0 (0%) | 7 (100%) | 0 (0%) | 0 (0%) | 12 (100%) | 0 (0%) | 0 (0%) | 11 (100%) | 0 (0%) | 0 (0%) | 187 (97.91%) | 3 (1.57%) | 1 (0.52%) |
| k13 | 515 | R | K* | 246 (99.19%) | 2 (0.81%) | 0 (0%) | 27 (100%) | 0 (0%) | 0 (0%) | 7 (100%) | 0 (0%) | 0 (0%) | 12 (100%) | 0 (0%) | 0 (0%) | 11 (100%) | 0 (0%) | 0 (0%) | 189 (98.95%) | 2 (1.05%) | 0 (0%) |
| k13 | 574 | P | L** | 260 (98.86%) | 1 (0.38%) | 2 (0.76%) | 28 (100%) | 0 (0%) | 0 (0%) | 7 (100%) | 0 (0%) | 0 (0%) | 13 (100%) | 0 (0%) | 0 (0%) | 12 (100%) | 0 (0%) | 0 (0%) | 200 (98.52%) | 1 (0.49%) | 2 (0.99%) |
| k13 | 622 | R | T | 248 (96.12%) | 5 (1.94%) | 5 (1.94%) | 27 (100%) | 0 (0%) | 0 (0%) | 6 (100%) | 0 (0%) | 0 (0%) | 12 (92.31%) | 0 (0%) | 1 (7.69%) | 12 (100%) | 0 (0%) | 0 (0%) | 191 (95.5%) | 5 (2.5%) | 4 (2%) |
| k13 | 675 | A | V** | 254 (98.45%) | 2 (0.78%) | 2 (0.78%) | 27 (100%) | 0 (0%) | 0 (0%) | 6 (100%) | 0 (0%) | 0 (0%) | 13 (100%) | 0 (0%) | 0 (0%) | 12 (100%) | 0 (0%) | 0 (0%) | 196 (98%) | 2 (1%) | 2 (1%) |
| mdr1 | 86 | N | Y | 254 (99.61%) | 0 (0%) | 1 (0.39%) | 27 (100%) | 0 (0%) | 0 (0%) | 5 (100%) | 0 (0%) | 0 (0%) | 12 (92.31%) | 0 (0%) | 1 (7.69%) | 10 (100%) | 0 (0%) | 0 (0%) | 200 (100%) | 0 (0%) | 0 (0%) |
| mdr1 | 182 | G | G | 255 (98.46%) | 1 (0.39%) | 3 (1.16%) | 27 (100%) | 0 (0%) | 0 (0%) | 7 (100%) | 0 (0%) | 0 (0%) | 13 (100%) | 0 (0%) | 0 (0%) | 11 (100%) | 0 (0%) | 0 (0%) | 197 (98.01%) | 1 (0.5%) | 3 (1.49%) |
| mdr1 | 184 | Y | F | 120 (46.33%) | 75 (28.96%) | 64 (24.71%) | 12 (44.44%) | 4 (14.81%) | 11 (40.74%) | 5 (71.43%) | 0 (0%) | 2 (28.57%) | 8 (61.54%) | 1 (7.69%) | 4 (30.77%) | 5 (45.45%) | 3 (27.27%) | 3 (27.27%) | 90 (44.78%) | 67 (33.33%) | 44 (21.89%) |
| mdr1 | 1246 | D | Y | 255 (98.08%) | 1 (0.38%) | 4 (1.54%) | 27 (100%) | 0 (0%) | 0 (0%) | 7 (100%) | 0 (0%) | 0 (0%) | 12 (92.31%) | 0 (0%) | 1 (7.69%) | 12 (100%) | 0 (0%) | 0 (0%) | 197 (98.01%) | 1 (0.5%) | 3 (1.49%) |
| mdr2 | 492 | I | V | 123 (47.49%) | 70 (27.03%) | 66 (25.48%) | 11 (40.74%) | 6 (22.22%) | 10 (37.04%) | 4 (66.67%) | 1 (16.67%) | 1 (16.67%) | 5 (38.46%) | 3 (23.08%) | 5 (38.46%) | 3 (25%) | 6 (50%) | 3 (25%) | 100 (49.75%) | 54 (26.87%) | 47 (23.38%) |

**Table S7. Proportion of samples carrying a haplotype of interest**

Haplotypes were reconstructed for samples with at most 1 amplicon with mixed genotype within the haplotype

| Genes | Haplotypes | N | % |
|---|---|---|---|
| mdr1 (86,184,1246) | NFD | 74 | 28.4% |
| | NFD+NYD | 58 | 22.2% |
| | NYD | 117 | 44.8% |
| | NYY | 1 | 0.4% |
| | Undetermined | 11 | 4.2% |
| dhfr (51,59,164) + dhps (436,437,540,581) | ICNSGEA | 1 | 0.4% |
| | ICNSGEA + ICNSGKA | 1 | 0.4% |
| | ICNSGKA | 3 | 1.1% |
| | ICNSGEA + IRNSGEA | 3 | 1.1% |
| | ICNSGKA+IRNSGKA | 7 | 2.7% |
| | IRNAGKA | 4 | 1.5% |
| | IRNAGKA + IRNSGKA | 1 | 0.4% |
| | IRNSGEA | 147 | 55.7% |
| | IRNSGEA +IRNSGEG | 6 | 2.3% |
| | IRNSGEA+IRNSGKA | 21 | 8.0% |
| | IRNSGEG | 17 | 6.4% |
| | IRNSGKA | 27 | 10.2% |
| | IRNSGEA+NRNSGEA | 3 | 1.1% |
| | IRNSGKA+NRNSGKA | 1 | 0.4% |
| | NRNSGEA | 1 | 0.4% |
| | NRNSGEG | 1 | 0.4% |
| | NRNSGKA | 1 | 0.4% |
| | Undetermined | 19 | 7.2% |

* Quintuple mutation haplotype: IRNSGEA; Sextuple mutation haplotype: IRNSGEG

**Table S8. Allele frequencies for minor alleles, adjusted by the complexity of infection of each sample.**

Synonymous mutations are denoted in lowercase. 95% confidence intervals shown in parenthesis.

| Gene | Codon | Allele | AlleleFrequency |
|---|---|---|---|
| arps10 | 127 | L | 0.002 (0.001-0.009) |
| crt | 74 | I | 0.006 (0.002-0.016) |
| crt | 75 | E | 0.006 (0.002-0.016) |
| crt | 76 | T | 0.006 (0.002-0.016) |
| dhfr | 108 | N | 0.999 (0.984-0.999) |
| dhfr | 51 | I | 0.955 (0.913-0.981) |
| dhfr | 59 | R | 0.896 (0.846-0.936) |
| dhps | 436 | A | 0.021 (0.011-0.036) |
| dhps | 437 | G | 0.999 (0.984-0.999) |
| dhps | 540 | E | 0.622 (0.566-0.676) |
| dhps | 581 | G | 0.059 (0.041-0.083) |
| k13 | 441 | L | 0.192 (0.157-0.23) |
| k13 | 479 | I | 0.004 (0.001-0.012) |
| k13 | 504 | a | 0.008 (0.003-0.019) |
| k13 | 515 | K | 0.004 (0.001-0.012) |
| k13 | 574 | L | 0.006 (0.002-0.016) |
| k13 | 578 | S | 0.002 (0.001-0.009) |
| k13 | 596 | V | 0.002 (0.001-0.009) |
| k13 | 622 | T | 0.021 (0.011-0.036) |
| k13 | 667 | A | 0.004 (0.001-0.012) |
| k13 | 675 | V | 0.006 (0.002-0.016) |
| mdr1 | 1246 | Y | 0.011 (0.004-0.022) |
| mdr1 | 182 | g | 0.008 (0.003-0.019) |
| mdr1 | 184 | F | 0.348 (0.302-0.396) |
| mdr1 | 86 | Y | 0.002 (0.001-0.009) |
| mdr2 | 492 | V | 0.347 (0.301-0.395) |
